# Classification and Predictors of Right Ventricular Functional Recovery in Pulmonary Arterial Hypertension

**DOI:** 10.1101/2023.02.15.23285974

**Authors:** Franz P. Rischard, Roberto J. Bernardo, Rebecca R. Vanderpool, Deborah H. Kwon, Tushar Acharya, Margaret M Park, Austin Katrynuik, Michael Insel, Saad Kubba, Roberto Badagliacca, A Brett Larive, Robert Naeije, Joe G.N. Garcia, Gerald J Beck, Serpil C Erzurum, Robert P Frantz, Paul M Hassoun, Anna R Hemnes, Nicholas S Hill, Evelyn M Horn, Jane A Leopold, Erika B. Rosenzweig, W.H. Wilson Tang, Jennifer D. Wilcox

## Abstract

**Background:** Normative changes in right ventricular (RV) structure and function have not been characterized in the context of treatment-associated functional recovery (RVFnRec). The aim of this study is to assess the clinical relevance of a proposed RVFnRec definition.

**Methods:** We evaluated 63 incident patients with PAH by right heart catheterization and cardiac MRI (CMR) at diagnosis and CMR and invasive cardiopulmonary exercise (CPET) following treatment (∼11 months). Sex, age, race/ethnicity matched healthy control subjects (n=62) with one-time CMR and non-invasive CPET were recruited from the PVDOMICS project. We examined therapeutic CMR changes relative to the evidence-based peak oxygen consumption (VO2_peak_)>15mL/kg/min to define RVFnRec by receiver operating curve analysis. Afterload was measured in the as mean pulmonary artery pressure, resistance, compliance, and elastance.

**Results:** A drop in RV end-diastolic volume of -15 mL best defined RVFnRec (AUC 0.87, P=0.0001) and neared upper 95% CI RVEDV of controls. 22/63 (35%) of subjects met this cutoff which was reinforced by freedom from clinical worsening, RVFnRec 1/21 (5%) versus no RVFnRec 17/42, 40%, (log rank P=0.006). A therapy-associated increase of 0.8 mL/mmHg in compliance had the best predictive value of RVFnRec (AUC 0.76, CI 0.64-0.88, P=0.001). RVFnRec subjects had greater increases in stroke volume, and cardiac output at exercise.

**Conclusions:** RVFnRec defined by RVEDV therapeutic decrease of -15mL predicts exercise capacity, freedom from clinical worsening, and nears normalization. A therapeutic improvement of compliance is superior to other measures of afterload in predicting RVFnRec. RVFnRec is also associated with increased RV output reserve at exercise.

**Clinical Perspective:** *What is new?:* Right ventricular functional recovery (RVFnRec) represents a novel endpoint of therapeutic success in PAH. We define RVFnRec as treatment associated normative RV changes related to function (peak oxygen consumption). Normative RV imaging changes are compared to a well phenotyped age, sex, and race/ethnicity matched healthy control cohort from the PVDOMICS project. Previous studies have focused on RV ejection fraction improvements. However, we show that changes in RVEDV are perhaps more important in that improvements in LV function also occur. Lastly, RVFnRec is best predicted by improvements in pulmonary artery compliance versus pulmonary vascular resistance, a more often cited metric of RV afterload.

*What are the clinical implications?:* RVFnRec represents a potential non-invasive assessment of clinical improvement and therapeutic response. Clinicians with access to cardiac MRI can obtain a limited scan (i.e., ventricular volumes) before and after treatment. Future study should examine echocardiographic correlates of RVFnRec.

## Introduction

Adaptation of the right ventricle (RV) to high afterload is the primary determinant of a patient’s symptoms and survival in PAH, making it a focus of research by expert working groups^1,2^. As afterload increases in PAH, the RV remodels with hypertrophy resulting in adaptive maintenance of cardiac output. Eventually, maladaptive processes occur such as myocardial interstitial fibrosis, detrimental myocyte intracellular and molecular changes^3,4^, and alterations in coronary flow^5^ leading to impaired systolic and diastolic function. When RV afterload is acutely reversed, for example with lung transplantation, there is often complete reversibility of these RV maladaptive changes^6^. In the pre-combination therapy era, the magnitude of afterload lowering was not large enough to appreciate substantial changes in RV structure^7^. However, combination therapy results in significant improvements in afterload^8^ making RV “reverse remodeling” a realistic therapeutic goal.

Reverse remodeling of the left ventricle (LV) is characterized by “normative” (returning toward normal structure and function) changes. But, also the left ventricular, cellular, and molecular processes associated with reverse remodeling have been well described^9^. Relative to the LV, characterization of RV reverse remodeling is in its infancy. Current definitions^7^ have relied upon the relationship to freedom from clinical worsening^10^ but moreover, they lack a substantive pathophysiological link of the consequences of pulmonary vascular load on RV function. In the absence of longitudinal RV cellular and molecular data, a formal definition of RV reverse remodeling is premature. However, RV functional recovery (RVFnRec) can be characterized by therapeutically associated “normative” imaging changes paired with peak exercise data (VO2_peak_) indicative of the RV stress response. To date, studies have not included a well-matched control group, so characterization of “normative” change is unknown. In addition, most studies thus far have used echocardiography to describe RVFnRec, whereas cardiac MRI (CMR) may be more sensitive to treatment-related changes^11^.

It is generally accepted that a large drop in afterload is required for RV reverse remodeling (or RVFnRec), but the most affected parameter of afterload is unknown^7^. Both steady (i.e., pulmonary vascular resistance, PVR) and pulsatile (i.e., pulmonary arterial compliance, Ca) components are important contributors to afterload in the pulmonary circulation^12^. Effective pulmonary arterial elastance (Ea), a composite measurement of total PVR and Ca, is also proposed as a more comprehensive way of representing afterload^13^. Lastly, parameters obtained during exercise such as alpha (α) distensibility, which describes the curvilinearity of the PA pressure-flow relationship, may give us insight into pulmonary vascular reserve/recruitment and the state of the distal vasculature^14^. No study has yet investigated these parameters longitudinally from PAH diagnosis to determine their impact on reverse remodeling.

In the present study, we aim to systematically define RVFnRec by CMR in a group of treatment naïve PAH patients relative to a VO2_peak_ threshold and clinical outcome. We evaluate the degree of normative RV therapeutic changes relative to matched healthy controls. We further assess the best parameter of RV afterload for predicting RVFnRec. Lastly, we examine if afterload and RV output reserve at exercise add to our understanding of RVFnRec.

## Methods

### Subjects

Sixty-three incident treatment naive patients with World Symposium of PH (WSPH) Group I PAH gave an informed consent to participate in the study, which was approved by the institutional review board at the University of Arizona (IRB #1100000621). The subjects enrolled as part of a prospective protocolized study which is open to all incident patients (See **supplemental methods**). The diagnosis of PAH was established by dedicated PAH providers with invasive confirmation by right heart catheterization based on updated guidelines^15^. Patients were included if they had right heart catheterization <u>and</u> CMR at baseline (pre-treatment) and invasive cardiopulmonary exercise testing (iCPET) <u>and</u> CMR at > 6 months of follow-up. All patients were placed on PAH goal-directed therapy per current guidelines at the time. See **supplemental methods** for details on therapeutic strategy. Subjects were categorized by therapeutic strategy based on the European Respiratory Society/European Society of Cardiology (ERS/ESC) guidelines of 2015 at which goal-directed mono or sequential therapy was replaced by up-front combination therapy^16^. We selected a cohort of patients all placed on parenteral treprostinil (± oral therapy) to optimize the possibility of seeing large changes in afterload (and thus, RVFnRec)^17^. Ninety-six healthy control subjects were recruited for the NHLBI Redefining Pulmonary Hypertension through Pulmonary Vascular Disease Phenomics (PVDOMICS)^18^. These subjects underwent extensive testing including CMR and non-invasive CPET at a single time point. Sixty-two age, sex, and race/ethnicity matched controls from across all clinical sites were used for this analysis with steering committee approval. The United States Registry to Evaluate Early and Long-Term PAH Disease Management (REVEAL) score 2.0 score^19^ was calculated and presented at baseline and follow-up for the PAH subjects.

### Cardiac MRI

All image analysis was performed using CVI42, Circle Cardiovascular Imaging® by TA (at Arizona) and DK (at the PVDOMICS Core, the Cleveland Clinic), expert cardiac MRI cardiologists with >10 years’ experience in CMR. Fifty subjects with overlap (between PVDOMICS and the UA registry) were analyzed by both TA <u>and</u> DK and used to assess inter-reader variability in RV volumes (see **supplemental methods**). Cardiac MRI imaging was performed as described previously^20^. Briefly, standard volumetric measurements were made from short axis cine projections for CMR. Contour smoothing was done to include trabeculations in end-systole (ESV) and diastole (EDV). Stroke volume (SV) was calculated as EDV – ESV and RVEF as [(SV/EDV) x 100]. Additional methods are available in the online supplement.

### Right Heart Catheterization and Invasive Cardiopulmonary Exercise Testing

After study inclusion, a pulmonary artery catheter was advanced via the antecubital vein for measurements of pulmonary artery pressure (systolic, mean, and diastolic PAP), RV pressure, right atrial pressure (RAP), wedged PAP (PCWP) and cardiac output (direct Fick). PVR is calculated as mPAP-PCWP divided by direct Fick cardiac output (C.O.) and expressed as WU (mmHg*min*L^-1^) or mmHg*sec*mL^-1^ where indicated. Ca was calculated as stroke volume (SV) from direct Fick C.O. divided by PA pulse pressure and expressed as mL*mmHg^-1^. Effective pulmonary arterial elastance (Ea) was calculated as sRVP/SV ^21^ expressed as mmHg*mL^-1^. Distensibility (α), expressed as % change in vessel diameter/mmHg distending pressure, was calculated by methods previously described ^14,22^. All stages of a single subject’s mPAP, PCWP, and C.O. were used to calculate a single α value. RC time, the product of PVR and Ca, was expressed as seconds.

Our comprehensive resting and exercise catheterization protocol has been previously published^23^. Briefly, after obtaining supine resting measurements, the patient was placed in full upright position with an electronic fluoroscopy chair. Fluoroscopy was used to re-zero at left atrial level. A cycle ergometer was positioned below the patient. The patient then proceeded with exercise at predetermined workload based on their level of dyspnea ^23^ for steady-state two-minute stages until respiratory exchange ratio (RER) ∼ 1.1. PAP, PCWP, RAP, arterial and venous O2 content (CaO2, CvO2) were obtained during the last 30 seconds of each stage. Metabolic cart analysis (Vyaire Medical™, Mettawa, IL) was used for simultaneous collection gas exchange and lung volume. Hemodynamics presented were averaged over 3 respiratory cycles in accordance with current guidelines at rest and exercise^24^. The control patients underwent an identical protocol in the non-invasive exercise lab (without a catheter in place).

### Definition of RV Functional Recovery

Since exercise VO2_peak_ is physiologically meaningful (dependent on RV function) and prognostic, the guideline^25^ and evidence (clinical worsening) confirmed^26^ cutoff of >15 mL/kg/min was chosen to identify potential CMR cutoffs indicative of RVFnRec. Exercise data is presented only for the follow-up visit as many patients (N=55 without iCPET, N=8 with iCPET) were not exercised at baseline due to safety considerations. We examined therapeutically associated change in RV size (RVESV and EDV, % change RV volumes) and function (RVEF) relative to this VO2_peak_ cutoff at follow-up. The Youden index was used to define the best cutoffs in RV volumes (absolute and relative change from baseline) and RVEF (change from baseline). Receiver operating characteristic (ROC) curves are used to display optimal cut-offs. Resulting RVFnRec groups were then examined relative to freedom from clinical worsening. **Clinical worsening** was defined as death, transplant, or hospitalization. Freedom from clinical worsening was evaluated using Kaplan-Meier Plots and the log-rank test. A similar procedure (Youden index and ROC analysis) was used to define the best cutoffs for mPAP, Ca, PVR, Ea, and α at follow-up and change from baseline to follow-up for predicting RVFnRec.

### Statistical Analysis

Continuous data are expressed as mean ± standard deviation or median [25,75 percentile]. Categorical data are expressed as counts and percentages. The number of PAH subjects was chosen based primarily on data availability given strict inclusion/exclusion criteria. However, based on previously published data in prostacyclin treated patients, we would expect 100% power to detect a mean difference of 7±5.4 WU PVR difference between RVFnRec and no RVFnRec groups (R.B. personal communication, 9/25/22 based on ref.^17^. Kruskal-Wallis test for continuous and ordinal variables or Fisher exact test for categorical variables were used to test baseline differences. Normality was assessed with the Shapiro-Wilk test. Longitudinal differences were evaluated with Wilcoxon signed-rank (within group) or repeated measures analysis of variance (ANOVA)(between group). Variables were log-transformed in cases of non-normal distribution. Linear or non-linear regression was conducted where appropriate after testing assumptions. Binary logistic regression was done to assess the relationship of treatment type with RVFnRec. Statistical analyses were performed using SPSS software (version 28.0, IBM, Armonk, NY) and SigmaPlot (version 14.5, Systat©, San Jose, CA). Statistical tests were 2-sided, and a p-value <0.05 was considered statistically significant. P values and 95% CIs presented in this report have not been adjusted for multiplicity, and therefore, inferences drawn from these statistics may not be reproducible.

## Results

### Patient Characteristics

Sixty-three treatment naïve PAH and 62 control subjects were prospectively enrolled. Subjects were predominantly female and PAH subjects were majority idiopathic (**Table 1**). At enrollment, the subjects had very advanced PAH, characterized by low cardiac index and significantly elevated afterload. CMR demonstrated nearly twice the RVEDVI and depressed RVEF relative to controls. Therapeutic improvements in RVEF were accompanied by improvements in 6-minute walk, PVR, pulmonary compliance, cardiac output, and BNP (**Table 1**).

**Table 1.** Demographics and clinical data, cardiac MRI, and resting hemodynamics by assessment time by PAH or control cohorts. Values are mean ± SD, median [P25,P75], or N (%). BMI, body mass index; CHD, congenital heart disease; CTD, connective tissue disease; HIV, human immunodeficiency virus; iPAH, idiopathic pulmonary arterial hypertension; PoPH, portopulmonary hypertension; BNP, brain natriuretic peptide; MRI, magnetic resonance imaging; RVESVI, right ventricular end-systolic volume index; RVEDVI, right ventricular end-diastolic volume index; RVEF, right ventricular ejection fraction; PAP, pulmonary artery pressure; PCWP, pulmonary capillary wedge pressure; RAP, right atrial pressure; PVR, pulmonary vascular resistance. P values calculated as follows: Kruskal-Wallis test for continuous and ordinal variables or Wilcoxon signed-rank non-parametric test for related samples, Fisher exact test for categorical variables. *Significant difference at level <0.05 within PAH follow-up versus baseline; † P<0.05 control versus follow-up 1 PAH.

|  |  | PAH (n=63) |  | Control (n=62) |
| --- | --- | --- | --- | --- |
|  |  | Baseline | Follow-up |  |
| Age (years) |  | 51.2±12.7 |  | 50.0±14.02 |
| Sex (N/% Female) |  | 51 (81) |  | 49 (79) |
| BMI (kg/m<sup>2</sup>) |  | 31.0±8.3 |  | **27.3±5.3†** |
| Race (N/% White) |  | 57 (90) |  | 61 (98) |
| Ethnicity (N/% Hispanic) |  | 18 (28) |  | 14 (23) |
|  | 6 Minute Walk Distance (M) | 284[115,362] | **355[204,434]\*** | **547[479,600]†** |
|  | BNP (pg/dL) | 503[127.5,852.5] | **70[27,155]\*** |  |
| Cardiac MRI | RVEDVI (mL/m<sup>2</sup>) | 110.1[92.2,136.1] | 100.8[80.7,128. | **69.8[59.2,78.8]†** |
|  | RVESVI (mL/m<sup>2</sup>) | 77.6[60,103.8] | **63.6[51.9,85.1]\*** | **29.9[23.8,36.1]†** |
|  | RVEF (%) | 27.4±9.3 | **35.0±9.8\*** | **57.12±7.29†** |
|  | RV Mass (gm/m<sup>2</sup>) | 46.1±15.2 | 45.0±22.8 | **10.8±3.4†** |
|  | RV Mass/EDV (gm/mL) | 0.41±0.12 | 0.41±0.14 | 0.17±0.05 |
| Hemodynamics | RAP (mmHg) | 11±7 | **4±4\*** |  |
|  | mPAP (mmHg) | 55.4±10.4 | **39.2±9.5\*** |  |
|  | PCWP (mmHg) | 9.2±4.4 | **7.3±3.1\*** |  |
|  | Cardiac Index (L/min/m<sup>2</sup>) | 2.02±0.63 | **2.9±0.99\*** |  |
|  | PVR (WU) | 11.8[9.45,15.9] | **6.1[4.79,7.98]\*** |  |
|  | Compliance (mL/mmHg) | 0.99[0.77,1.35] | **1.74[1.30,2.33]\*** |  |
|  | PA Elastance (mmHg/mL) | 1.52[1.19,1.99] | **0.89[0.70,1.28]\*** |  |

### Right Ventricular Functional Recovery

As demonstrated in **Figure 1A-B**, VO2_peak_ was correlated with RVEDV and RVEF. 22/63 (35%) subjects had VO2_peak_>15mL/kg/min with 20 (91% of the 22) having a change in RVEDV and RVEF meeting the cutoff defined by the highest Youden index (0.74 at - 15mL and 0.51 at 4% change, respectively) while 2 (9%) met only one of these cutoffs. Of the 41/63 (65%) of subjects with VO2_peak_≤15mL/kg/min, 22 (54% of the 41) were below both cutoffs, 15 (37%) had RVEF above the 4% change but below the RVEDV cutoff, whereas 4 (9%) met both the RVEF and RVEDV cutoffs. ROC analysis (**Figure 1C**) confirmed changes in RVEDV were superior to changes in RVEF in the prediction of VO2_peak_. Indexing RVEDV to BSA (RVEDVI) was not more predictive than RVEDV (Youden 0.65 at -10.4 mL/m^2^). Change in RVESV and relative change in RVESV and EDV were not more predictive than change in RVEDV (**Table E1**). All PAH subjects who achieved the high VO2_peak_ cutoff demonstrated improvements in both RVESV and RVEDV, whereas some subjects below the VO2_peak_ cutoff had improvements in RVESV alone (**Figure E1**).

**Figure 1.**
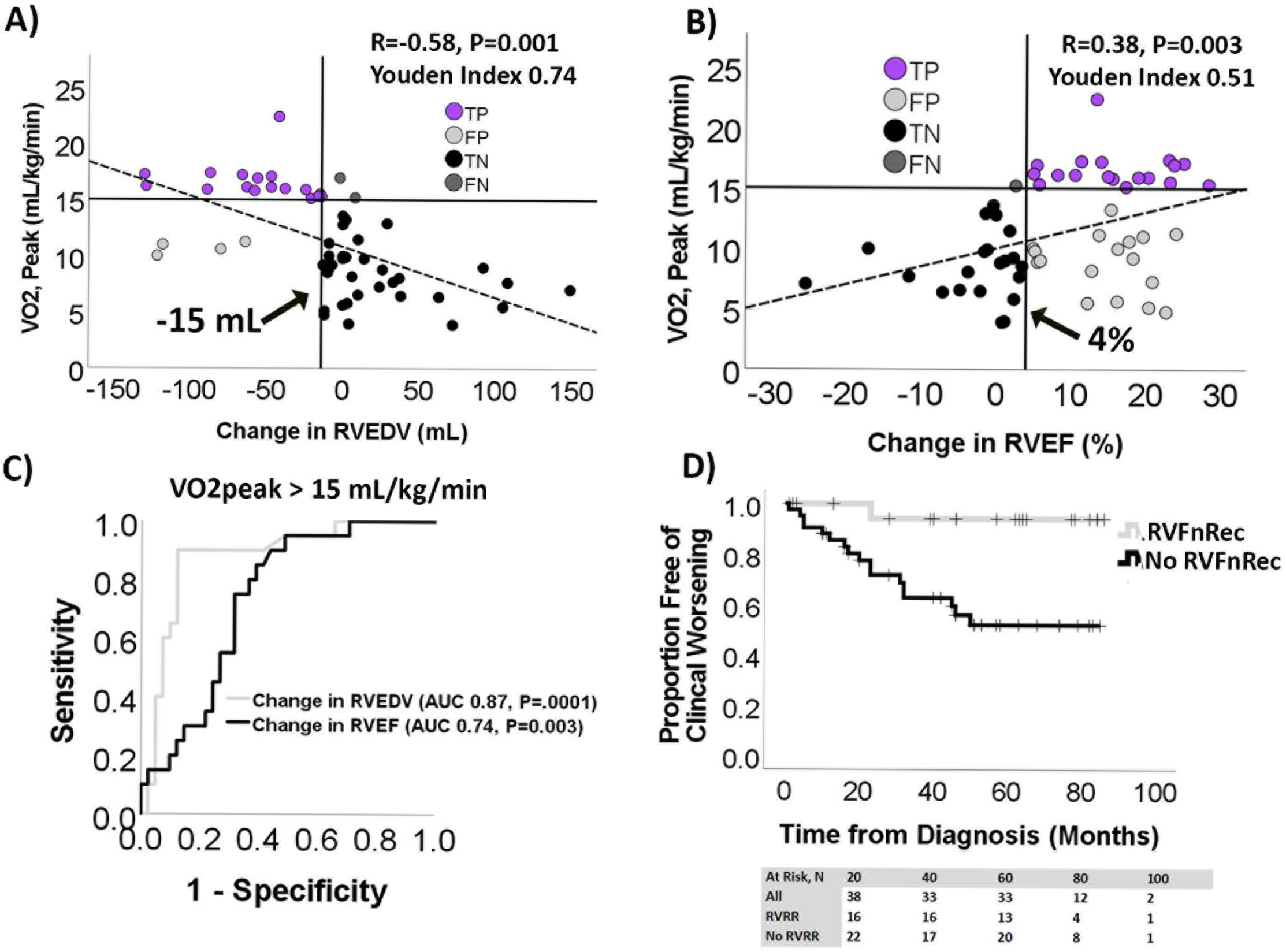
Defining right ventricular functional recovery. A change in RV end-diastolic volume (RVEDV) **(A)** and RV ejection fraction (RVEF) **(B)** from baseline to follow-up both correlated with peak oxygen consumption, VO2_peak_, at follow-up. However, a change in RVEDV better discriminated high exercise capacity (VO2_peak_ >15mL/kg/min) than did a change in RV ejection fraction (RVEF). Vertical solid lines indicate best Youden index of 0.72 at an RVEDV of -15mL or 0.51 for RVEF at +4%. Based on these cutoffs, the number of true positives (TP) and true (TN) and false (FN) negatives were similar in RVEDV and RVEF. However, the number of false positives (FP) was higher for RVEF. **C)** Receiver operating characteristic (ROC) analysis predicting VO2_peak_>15 mL/kg/min shows a change in RVEDV of -15mL is more predictive than RVEF +4mL as well. **D)** Kaplan-Meier plot of RVFnRec and no RVFnRec by clinical worsening shows higher freedom from clinical worsening in the RVFnRec group.

22/63 (35%) of subjects met criteria for RVFnRec using a drop of -15 mL in RVEDV. Using this definition of RVFnRec, the average length of follow up was 50±28 and 38±25 months for the RVFnRec and no RVFnRec groups, respectively. All-cause mortality was 1 (5%) and 10 (24%) and clinical worsening was 1 (5%) and 17 (41%) in the RVFnRec and no RVFnRec groups, respectively. As shown in **Figure 1D**, this definition of RVFnREc also resulted in a significant difference in time to clinical worsening, P=0.006. There were significantly more subjects in the RVFnRec group treated with up-front combination therapy versus the no RVFnRec group (50 versus 24%, P=0.01, **Table 2 and Figure E2**). Treatment approach differences were due in part to a difference between groups in time of enrollment (RVFnRec median enrollment date 4/2017 [3/2014-5/2021] versus no RVFnRec 5/2018 [11/2013-12/2021]). Up-front combination therapy was associated with odds-ratio of 10.4 (CI 1.9-56.6, P=0.007) of RVFnRec relative to goal-directed sequential therapy. There were significant improvements in 6-minute walk distance, REVEAL 2.0, RV volumes and EF in the RVFnRec group by follow-up compared to the no RVFnRec group (**Table 2 and 3)**. Although both groups had an increase in LVEDV, only the RVFnRec group had an improvement in LVEF (**Table 3**).

**Table 2.** Demographics and clinical data, multiparametric risk score, and standard resting hemodynamics by assessment time by RV functional recovery (RVFnRec) or no RV functional recovery (No RVFnRec) cohorts. Values are mean ± SD, median [P25,P75], or N (%). ADO, aldosterone; BMI, body mass index; CHD, congenital heart disease; CTD, connective tissue disease; HIV, human immunodeficiency virus; iPAH, idiopathic pulmonary arterial hypertension; PoPH, portopulmonary hypertension; REVEAL, United States Registry to Evaluate Early and Long-Term PAH Disease Management; ERS, European Respiratory Society; BNP, brain natriuretic peptide; MRI, magnetic resonance imaging; RVESVI, right ventricular end-systolic volume index; RVEDVI, right ventricular end-diastolic volume index; RVEF, right ventricular ejection fraction; PAP, pulmonary artery pressure; PCWP, pulmonary capillary wedge pressure; RAP, right atrial pressure; PVR, pulmonary vascular resistance; TRE, treprostinil. P values calculated as follows: Kruskal-Wallis test for continuous and ordinal variables or Wilcoxon signed-rank non-parametric test for related samples (within cohort) or repeated measures ANOVA (between cohort), Fisher exact test for categorical variables. * P<0.05 versus baseline within RVFnRec or no RVFnRec cohort; † P<0.05 between cohorts.

|  |  | RVFnRec (n=22) |  | No RVFnRec (n=41) |  |
| --- | --- | --- | --- | --- | --- |
|  |  | Baseline | Follow-Up | Baseline | Follow-Up |
| Age (years) |  | 54.1±9.4 |  | 49.7±14.0 |  |
| Sex (N/% Female) |  | 19 (91) |  | 32 (76) |  |
| BMI (kg/m<sup>2</sup>) |  | 31.4±10.5 |  | 30.9±7.1 |  |
| WSPH Category, N (%) | CHD | 1 (4.8) |  | 1 (2.4) |  |
|  | CTD | 4 (19.0) |  | 11 (26.2) |  |
|  | Drug/Toxin | 3 (14.3) |  | 8 (19.0) |  |
|  | HIV | 0 (0) |  | 1 (2.4) |  |
|  | iPAH | 12 (57.1) |  | 20 (47.6) |  |
|  | PoPH | 1 (4.8) |  | 1 (2.4) |  |
| Risk Scores, N (%) | REVEAL 2.0 Low Risk | 1 (4.8) | 20 (91.0) | 7 (16.7) | 20 (47.6) |
|  | REVEAL 2.0 Intermediate Risk | 7 (33.3) | 2 (9.0) | 12 (28.6) | 11 (26.2) |
|  | REVEAL 2.0 High Risk | 13 (61.9) | 0 (0) | 23 (54.8) | 11 (26.2) |
|  | 6 Minute Walk Distance (M) | 267±117 | **412±101\*†** | 252±129 | 274±141 |
|  | BNP (pg/dL) | 343[134,706.0] | **60[27,127]\*** | 423[128,853] | **76[28,177]\*** |
| Treatment, N (%) | Monotherapy TRE (prior to 2015) |  | 9 (41) |  | 12 (29.3) |
|  | Sequential TRE + Oral (prior to 2015) |  | **2 (9.1)†** |  | **19 (46.3)** |
|  | Up Front TRE + Oral (after 2015) |  | **11 (50)†** |  | **10 (24.4)** |
| Diuretics, N (%) | Loop Diuretics |  | 6 (27.3) |  | 12 (29.3) |
|  | Loop + ADO Antagonist |  | 2 (9.1) |  | 2 (4.9) |
|  | Loop + Metolazone |  | 1 (4.5) |  | 0 (0) |
| Hemodynamics | RAP (mmHg) | 11±6 | **3.0±3.0\*** | 11±7 | **5±4\*†** |
|  | mPAP (mmHg) | 56.5±11.4 | **34.7±8.4\*** | 54.8±10 | **41.5±9.4\*†** |
|  | PCWP (mmHg) | 9.4±4.2 | 5.4±3.2 | 9.1±4.6 | 6.7±3.0 |
|  | Cardiac Index (L/min/m<sup>2</sup>) | 1.98±0.7 | **3.27±1.2\*†** | 2.0±0.6 | **2.7±0.8\*** |

**Table 3.** Longitudinal Cardiac MRI features of the RV functional recovery (RVFnRec) and no recovery (No RVFnRec) cohorts. Values are mean ± SD, median [P25,P75], or N (%). RVEDVI, right ventricular end-diastolic volume index; RVESVI, right ventricular end-systolic volume index; RVEF, right ventricular ejection; RVMI, right ventricular mass index; RV M/V, right ventricular mass/volume; RV SV/ESV, right ventricular stroke volume/end-systolic volume; LVEDVI, left ventricular end-diastolic volume index; LVESVI, left ventricular end-systolic volume index; LVEF, left ventricular ejection-fraction; LVMI, left ventricular mass index; TR volume, tricuspid regurgitant volume; TR fraction, tricuspid regurgitant volume fraction (relative to stroke volume) fraction. P values calculated as follows: Wilcoxon signed-rank non-parametric test for related samples (within cohort) or repeated measures ANOVA (between cohort). * within cohort change from baseline to follow-up; † p value for between cohort change baseline to follow-up; ‡ p-value for between group relative change (as % baseline value) baseline to follow-up.

| RVFnRec (n=22) |  |  |  |  | No RVFnRec (n=41) |  |  |  |  |  |
| --- | --- | --- | --- | --- | --- | --- | --- | --- | --- | --- |
|  | Baseline | Follow-Up | Mean Change | Relative Change | Baseline | Follow-Up | Mean Change | Relative Change | P Value† | P Value‡ |
| **RVEDVI (mL/m2)** | 119.5[104.0,145.7] | **92.3[75.5,102.7]\*** | -34.5±24.6 | -25.5±14.1 | 105[87.5,131.8] | 114.0[88.2,159.0] | 8.7±26.5 | 9.0±26.8 | **<0.001** | **<0.001** |
| **RVESVI (mL/m2)** | 92.3[74.5,105.1] | **35.5[31.0,40.0]\*** | -37.6±18.2 | -39.6±13.3 | 71.2[57.4,101.9] | 73.2[57.5,123.8] | 1.6±25.1 | 6.2±37.7 | **<0.001** | **<0.001** |
| **RVEF (%)** | 26.6±6.7 | **43.0±7.4\*** | 15.7±7.1 | 55.7±35.9 | 27.7±10.3 | 32.0±8.7 | 4.4±11.0 | 32.4±62.8 | **<0.001** | 0.12 |
| **RVMI (gm/m2)** | 44.6[34.1,51.3] | 35.4[29.8,40.5] | -1.1±28.2 | -2.1±41.0 | 50.0[29.4,60.5] | 43.9[33.0,59.2] | -1.2±13.0 | 0.2±30.6 | 0.94 | 0.89 |
| **RV M/V (gm/mL)** | 0.4[0.3,0.52] | 0.37[0.33,0.45] | 0.1±0.1 | 22.0±33.7 | 0.42[0.3,0.6] | 0.34[0.30,0.50] | -0.04±0.1 | -7.2±22.8 | 0.06 | 0.07 |
| **RV SV/ESV** | 0.37±0.1 | **0.72±0.3\*** | 0.34±0.2 | 98.0±66.8 | 0.41±0.2 | 0.5±0.2 | 0.1±0.2 | 51.4±93.9 | **<0.001** | **0.04** |
| **LVEDVI (mL/m2)** | 47.5[44.8,59.7] | **66.3[51.5,75.9]\*** | 10.7±16.9 | 28.6±44.3 | 49.0[39.5,54.9] | **63.8[55.1,76.2]\*** | 16.8±15.4 | 35.4±32.4 | 0.22 | 0.56 |
| **LVESVI (mL/m2)** | 25.0[17.1,26.9] | 26.6[17.7,36.4] | 1.7±9.9 | 12.6±46.0 | 22.0[15.3,27.8] | **26.6[23.2,32.8]\*** | 8.3±9.3 | 46.8±52.4 | **0.02** | **<0.001** |
| **LVEF (%)** | 54.0[46.2,61.3] | **56.8[54.8,68.3]\*** | 7.2±9.9 | 15.6±19.9 | 54.0[46.9,67.3] | 57.1[53.3,60.8] | -1.3±11.5 | -0.2±19.8 | **0.01** | **0.01** |
| **LVMI (gm/m2)** | 36.8±6.1 | 42.4±26.3 | 22.0±5.1 | 62.3±13.2 | 45.0±16.9 | 42.3±18.7 | -3.5±19.8 | 1.1±52.0 | 0.12 | 0.15 |
| **TR volume (mL)** | 3.0[0,14.0] | 7.0[0,21.8] | 7.6±10.9 |  | 6.0[0,16] | 5.0[0,16.0] | -0.8±16.4 |  | 0.4 |  |
| **TR fraction (%)** | 0.0[0,24.6] | 9.8[0,33.6] | 10.1±18.5 |  | 5.0[0,32] | 8.2[0,27.7] | -2.5±21.4 |  | 0.2 |  |

As shown in **Figure 2A**, only the RVFnRec group demonstrated a therapeutic change in RVEDV that nears “normalization” of RVEDVI at the upper 95% confidence interval (CI) of healthy controls (90 mL/m2). -15 mL decrease in RVEDV identifies most of the patients included in this confidence interval. Although most of both the RVFnRec(20/22, 91%) and no RVFnRec (32/41, 78%) groups met the SVI lower 95% CI of controls (30 mL/m2), only the RVFnRec group neared the upper and lower 95% CI of controls for RVEDVI (11/22, 50% versus 10/41, 24%, respectively) and RVEF (8/22, 35%) versus 2/41, 5%, respectively) at follow-up (**Figure 2B**). Despite near normalization of EDV and RVEF among the RVFnRec group, relative wall thickness (RV mass/volume) remained approximately twice that of controls, RVFnRec 0.43±0.13 gm/mL versus control 0.21±0.05 gm/mL (**Figure 2C, Table 1 and 2**). **Table E2** compares RV volume and mass in PAH and control subjects with previously published control reference values.

**Figure 2.**
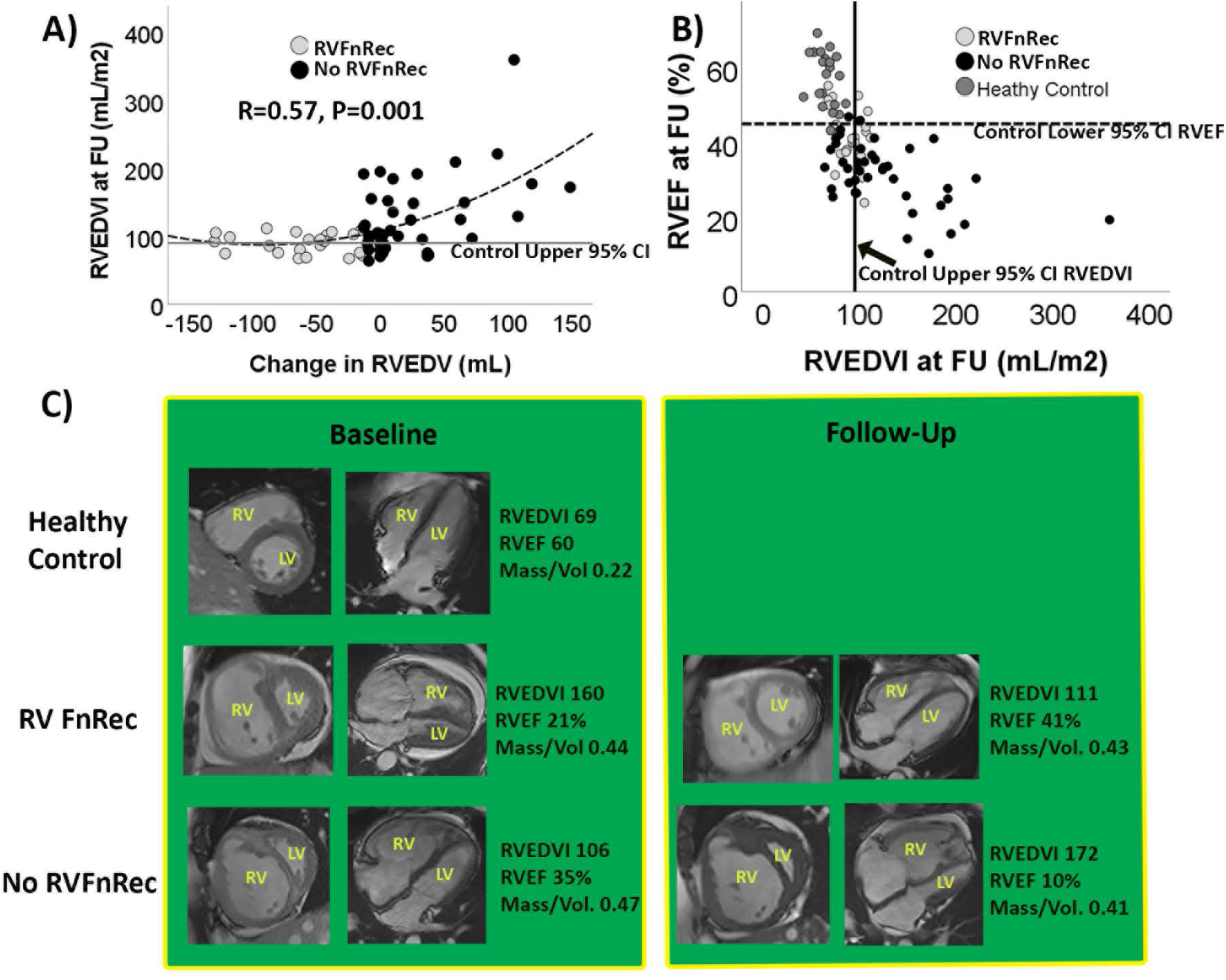
Near normalization of right ventricular volume in functional recovery. **A)** The treatment related change in RV end-diastolic volume (RVEDV) relative to RVEDV index at follow-up in the functional recovery (RVFnRec) and no RVFnRec subjects. Only RVFnRec subjects achieve a therapeutic change in RVEDV that nears normalization. **B)** RVEF and RVEDVI at follow-up in the RVFnRec, no RVFnRec, and healthy controls. Solid vertical line indicates the upper 95% CI and the dark horizontal line the lower 95% CI for RVEDV and RVEF in the control cohort, respectively. **C)** Treatment related changes in short (first column) and long (second column) axis cardiac MRI images at Baseline and Follow-up. Substantial, improvements in RV volume and ejection fraction are seen in the RVFnRec group. However, relative wall thickness (mass/volume) remained almost twice the healthy control value in both the RVFnRec and no RVFnRec groups.

### RV Afterload Parameters Associated with RV Functional Recovery

Patients with RVFnRec had larger absolute and relative (to baseline) drops in mPAP, and Ca, as well as relative changes in PVR, versus the no RVFnRec group by follow-up (**Table 4**). The absolute change in PVR and Ea and relative changes in Ea were not significant (**Table 4**). A greater proportion of RVFnRec than no RVFnRec subjects had moved to the “steep” portion of the PVR-Ca curve (**Figure 3A**) by follow-up. 9/63 (∼14%) of PAH subjects had near normalization of PVR (∼3WU) (**Figure 3A**). The accuracy of the change in the absolute value, change in relative value (as % baseline), and absolute value at follow-up were assessed as shown in **Figure 3B and E3A-B**. The value giving the best AUC 0.76 (95% CI 0.64-0.88, P<0.0001) was change in Ca at a cutoff of 0.8 mL/mmHg yielding a sensitivity of 85% and a specificity of 72% for RVFnRec. Changes in mPAP, PVR, and Ca were associated with changes in RVEDV (**Figure 3 C-E**) and RVEF (**Figure E3C-E**) with Ca being the most highly predictive.

**Table 4.**
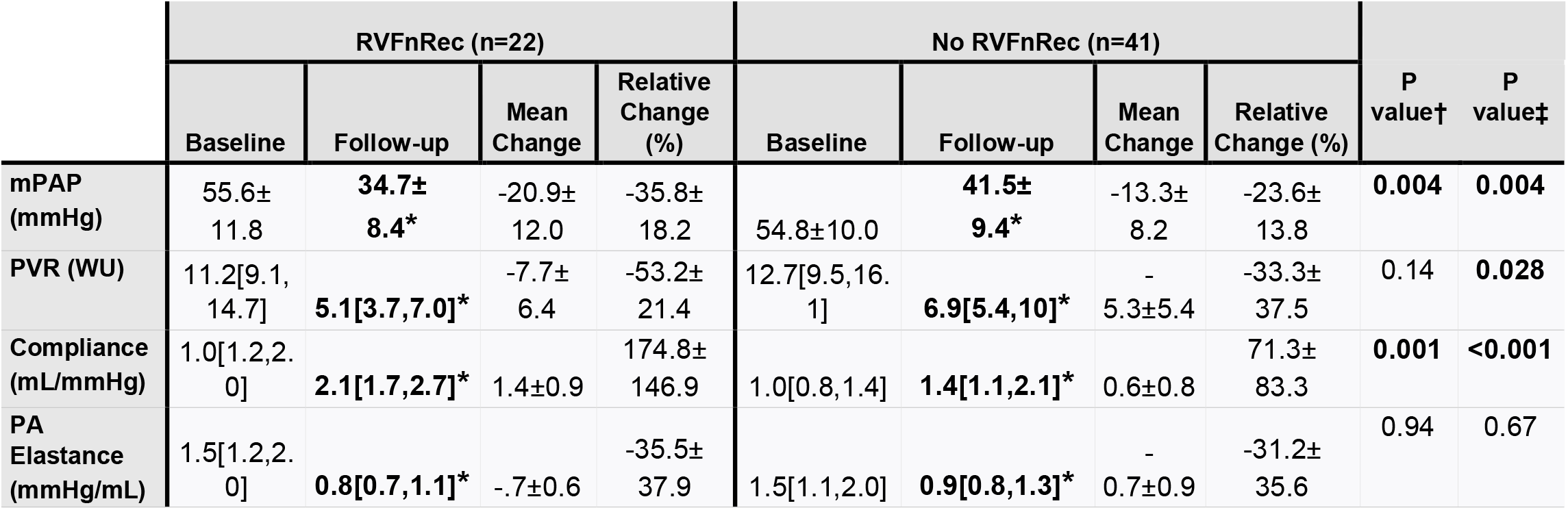
Longitudinal parameters of afterload in the RV functional recovery (RVFnRec) and no recovery (No RVFnRec) cohorts. Values are mean ± SD, median [P25,P75], or N (%). mPAP, mean pulmonary artery pressure; PVR, pulmonary vascular resistance; PA, pulmonary artery. P values calculated as follows: Wilcoxon signed-rank non-parametric test for related samples (within cohort) or repeated measures ANOVA (between cohort). * within cohort change from baseline to follow-up; † p value for between cohort mean change baseline to follow-up; ‡ p-value for between group relative change (as % baseline value) baseline to follow-up.

**Figure 3.**
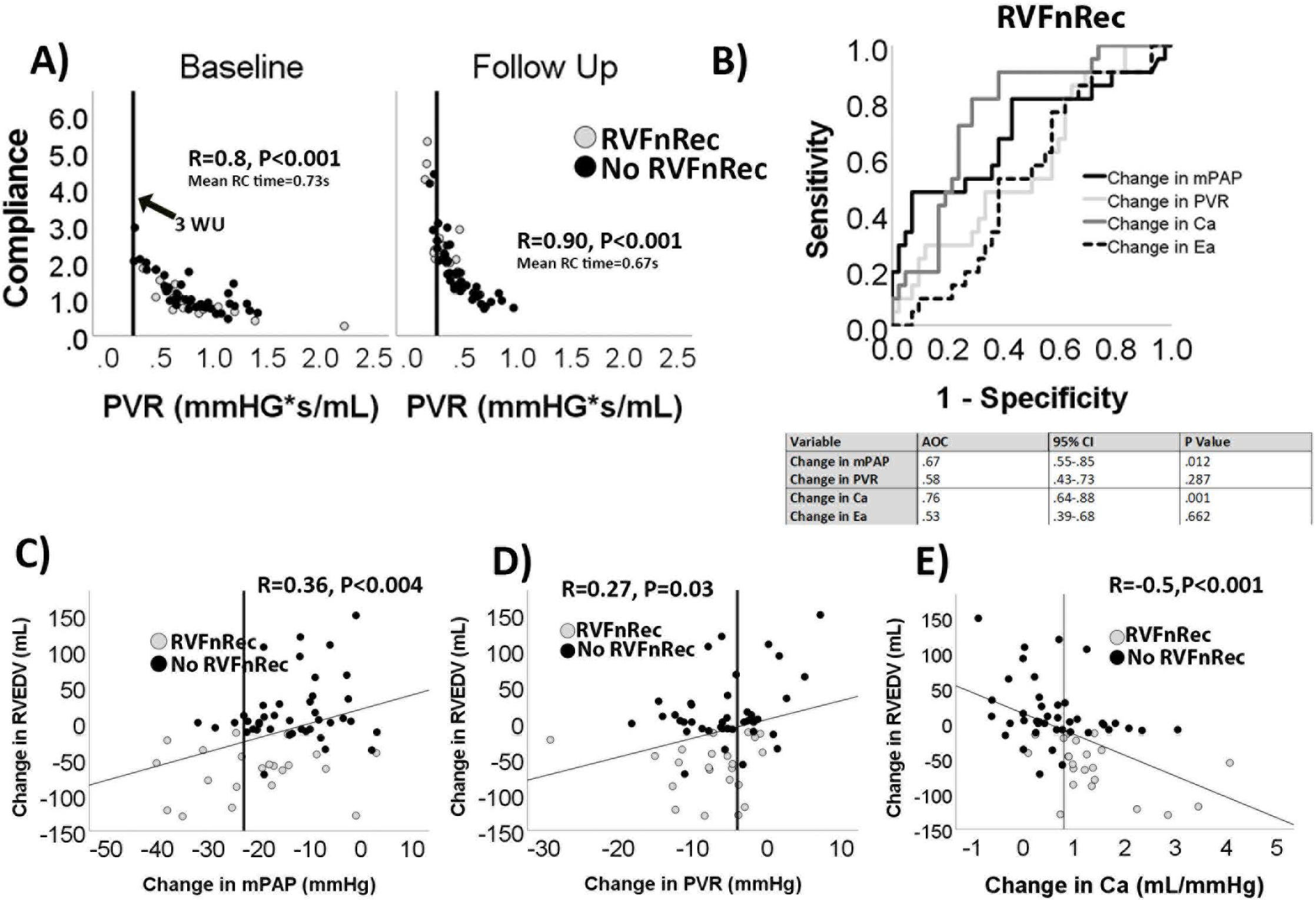
Afterload changes predicting right ventricular functional recovery. (**A**) non-linear relationship between pulmonary vascular resistance (PVR) and compliance (Ca) at baseline and follow-up. Most RVFnRec subjects had moved to the steep portion of the curve by follow-up. Solid line indicates an upper normal PVR of <3 WU. (**B**) receiver operating curve analysis of longitudinal changes between baseline and follow-up in mean pulmonary artery pressure (mPAP), PVR, Ca, and effective pulmonary elastance (Ea). Changes in Ca were the most closely related afterload parameter to changes in RV end-diastolic volume (RVEDV) (**C-E**). Vertical line indicates the highest (“best”) Youdin index cutoff for each afterload parameter.

### The Relationship of RV Functional Recovery to Exercise Afterload and RV Output Reserve

The RVFnRec subjects had higher VO2_peak_ predicted, mPAP, cardiac output, and stroke volume at exercise than did the no RVFnRec subjects at follow-up (**Figure 4 and Table 5)**. The VO2_peak_ was much lower than matched controls, however. Ventilatory efficiency index for carbon dioxide (Ve/VCO2) was lower (better) in the RVFnRec group. The mPAP-cardiac output relationship from rest to peak exercise, was similar in RVFnRec versus the no RVFnRec group (**Figure 4B**). α distensibility was also similar between RVFnRec and no RVFnRec groups (0.17±0.15 versus 0.19±0.3 %/mmHg, P=NS, respectively) and much lower than predicted normal values of 1.4 (range 0.77-2.34) %/mmHg^14^(**Figure 4D**). Using exercise afterload parameters did not improve the accuracy of resting values at follow-up to predict RVFnRec (**Figure E4**).

**Table 5.** RV functional recovery (RVFnRec) and No RV functional recovery (No RVFnRec) cohorts peak invasive cardiopulmonary exercise at follow-up and healthy control non-invasive peak exercise variables. Values are mean ± SD, median [P25,P75]. P values calculated from the Kruskal-Wallis test. RER, respiratory exchange ratio; VO2, peak oxygen consumption; RAP, right atrial pressure; mPAP, mean pulmonary artery pressure; PCWP, pulmonary capillary wedge pressure; PAO2, pulmonary arterial oxygen saturation; PVR, pulmonary vascular resistance; Ve/VCO2, minute ventilation/exhaled carbon dioxide; PetCO2, end-tidal CO2; O2pulse, oxygen pulse (VO2/heart rate), CaO2, arterial oxygen content; CvO2, pulmonary arterial oxygen content; Ca-vO2, arterio-venous oxygen content difference; α distensibility, distensibility of the pulmonary arterial system (calculated at exercise). *P<0.05 for RVFnRec versus No RVFnRec; †=P<0.05 for Health Control versus RVFnRec: ‡P<0.05 for Healthy Control versus No RVFnRec.

|  | RVFnRec (n=22) | No RVFnRec (n=41) | Healthy Control (n=62) |
| --- | --- | --- | --- |
| RER | 1.14±0.10 | 1.06±0.11 | 1.16±0.08†‡ |
| Work, (Watts) | 35[20,60] | 20[10,40] | 140[118,198]†‡ |
| VO<sub>2</sub> (mL/kg/min) | 15.1±3.2* | 8.9±3.0 | 23.73±7.7†‡ |
| VO<sub>2</sub><sub>predicted</sub> | 62±14* | 39±12 | 100±26†‡ |
| Heart Rate (bpm) | 115[104.0,120.0] | 110[98,121] | 157.0[146.0,166.0]†‡ |
| RAP (mmHg) | 7±4* | 11±7 |  |
| mPAP (mmHg) | 62.5[50.0,69.0] | 58.8[48.0,68.0] |  |
| PCWP (mmHg) | 10.6±4.9 | 10.8±4.1 |  |
| Cardiac Output (L/min) | 9.9[7.8,12.9]* | 8.3[6.0,10.3] |  |
| PAO<sub>2</sub><sub>sat</sub> (%) | 45±8 | 41±10 |  |
| PVR (WU) | 5.2[3.1,6.6] | 5.6[4.3,7.7] |  |
| Compliance (mL/mmHg) | 2.28±0.9 | 1.9±0.7 |  |
| Ve/VCO<sub>2</sub> at AT | 37.0[35.0,41.0]* | 42.0[39.0,48.2] | 29.6[27.3,32.2]†‡ |
| PetCO<sub>2</sub> at AT | 31.1±3.8* | 27.3±6.5 |  |
| O<sub>2</sub>pulse | 7.0[6.0,8.1] | 6.1[5.3,7.1] | 10.7[9.2,13.0]†‡ |
| CaO<sub>2</sub> (mL/dL) | 176.4±21.1 | 178.8±21.0 |  |
| CvO<sub>2</sub> (mL/dL) | 87.3±18.4 | 79.6±17.0 |  |
| Ca-vO<sub>2</sub> (mL/dL) | 90.4±18.1 | 87.9±30 |  |
| α distensibility (%/mmHg) | 0.914[0.00,0.231] | 0.130[0.268,0.290] |  |

**Figure 4.**
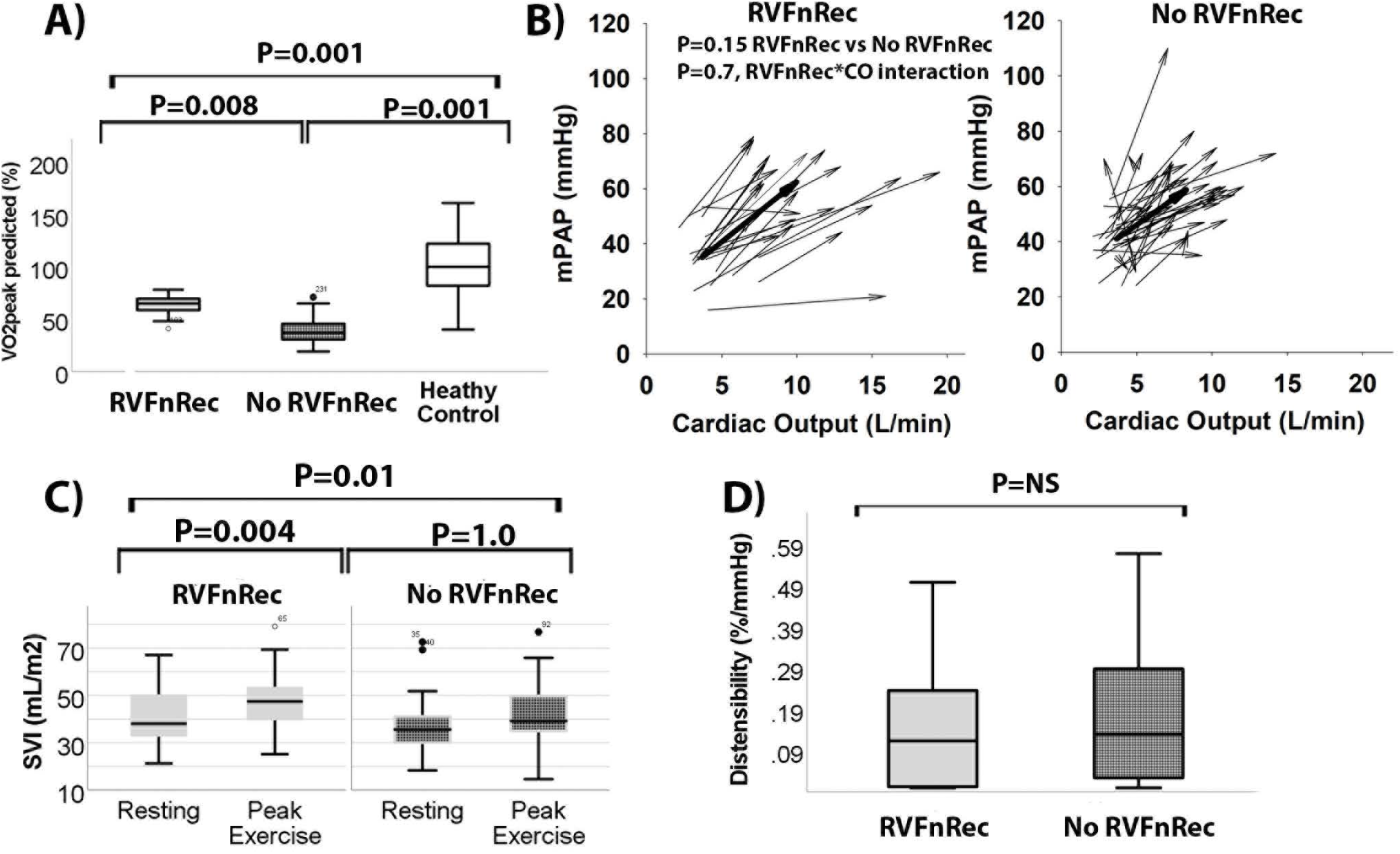
The physiological significance of right ventricular functional recovery through exercise. Peak predicted oxygen consumption was higher in RVFnRec than no RVFnRec but both were well below normal matched controls (**A**). The linear pressure-flow relationship of RVFnRec subjects and no RVFnRec subjects was similar (**B**). The arrowheads represent peak and the tails resting mPAP/cardiac output. The thick arrow represents the median vector in each group. Despite similar mPAP/cardiac output, the cardiac output at peak was higher (**B**) and the change in stroke volume index (SVI) (**C**) was higher in RVFnRec than that of no RVFnRec subjects. Pulmonary vascular α distensibility was similar between groups (**D**). Taken together (**B-D**) these findings indicate similar RV afterload at exercise but improved RV output reserve in the RVFnRec cohort.

## Discussion

This study is the first to demonstrate in treatment naïve PAH patients that 1) a -15 mL decrease in RVEDV is a useful CMR definition of RVFnRec associated with exercise capacity (VO2_peak_) and freedom from clinical worsening, 2) an increase in resting Ca of 0.8 mL/mmHg is superior to other afterload parameters in predicting RVFnRec, and 3) RVFnRec is associated with high exercise RV output reserve.

Although therapeutic improvements in RV size and function add independently prognostic information to well validated risk scores (such as REVEAL 2.0 and ERS risk score) in PAH^10,27^, the lack of a consensus definition of RV recovery hampers its use as a therapeutic goal and a clinical trial endpoint. As normalization of RV size and function would be an optimal goal, defining the limits of “normal” is challenging given reference populations have varied by age, sex, race/ethnicity, body mass index, and acquisition/reading methodology^28-30^. We have attempted to mitigate these challenges by enrolling a control group matched for age, sex, and race/ethnicity. Although many of our subjects met the CMR 95% confidence interval in the controls, normalization appears to be an unrealistic definition of RVFnRec. However, complete normalization while a laudable goal may be unnecessary for meaningful RV functional recovery. Unlike definitions of RV reverse remodeling which rely on the absence of clinical worsening alone^10^, we thought it important to examine RVFnRec by CMR cutoffs relating to clinical improvement and exercise capacity. The VO2_peak_ chosen at 15 mL/kg/min is dependent on cardiac output (RV functional reserve) in PAH^26^ and is derived from guidelines^25^ and evidence^26^. We confirmed the findings of others by showing changes in RVEDV are related to freedom from clinical worsening in the RVFnRec group.

There is consensus that RVEF is consistently related to mortality ^1^ and therapeutic improvements in RVEF are related to a drop in afterload ^10,31^. Studies enrolling subjects on oral monotherapy or sequential therapy result in minimal afterload reduction. In these studies, improvement in RVEF occurs by reductions in RVESV but with minimal change in RVEDV^27,31-33^. Early therapeutic changes in RVESV result from improvements in afterload and RV diastolic function^20^. But RV work efficiency does not improve (vis a vis improved RV-PA coupling) until there is a drop in RVEDV with larger afterload reduction (∼7WU or >45% drop in PVR)^34^. In fact, we demonstrate that a drop in both RVESV and RVEDV are highly associated with an acceptable (VO2_peak_ >15mL/kg/min) exercise capacity. We attempted to optimize the probability of large changes in afterload by employing an aggressive therapeutic strategy based on parenteral prostacyclin ^7,17^. A drop in RVEDV may signify a physiological shift from both heterometric (Frank-Starling) and homeometric (contractility) adaptation to maintain stroke volume to a more energetically optimized condition of homeometric adaptation alone^35^. Although improvements in RVEF from decreased RVESV represent a formidable short-term therapeutic goal, the increased myocardial efficiency and reduced wall stress accompanied by decreasing RVEDV should represent the ultimate long-term goal ^35^.

Clinical observation indicates that biventricular maladaptive remodeling in PH is reversible with rapid hemodynamic normalization such as lung transplantation^6^ and pulmonary thomboendarterectomy (PTE) for chronic thromboembolic PH (CTEPH)^36^. Even RV fibrosis appears reversible in experimental PAH when afterload is gradually but completely normalized^37^. We have shown for the first time that biventricular normative changes occur particularly in up-front prostacyclin with oral combination therapy resulting in a significant and gradual drop in afterload. Only patients with RVFnRec demonstrated large reductions in RV volume and improvements in RV and LVEF. Therefore, a reduction in RVEDV may be requisite to an improvement in the LV maladaptive changes, such as reduced LVEF^36^, seen with PAH and CTEPH. Although RVEDV shows near “normalization” relative to controls, normalization of RVEF and mass/volume was less common. RV relative wall thickness (mass/volume) was maintained more than double that of controls likely reflecting reduced but persistently high load. In this context, the present definition of RVFnRec is near the prognostically significant cut-point of 0.47 gm/mL^38^. The combination of RVFnRec and a high RV mass/volume appears physiologically significant during exercise where these subjects can maintain higher cardiac output and stroke volume (RV output reserve) as compared to those without RVFnRec despite similar PAP^39,40^.

As recently reviewed^7^, knowledge gaps exist on which afterload parameter predicts RVFnRec. Although the pulmonary impedance spectrum is the gold standard of afterload measurement, it requires simultaneous pressure and flow measurement in the frequency domain making it difficult to measure and interpret^12^. We have therefore chosen a comprehensive list of frequently measured afterload parameters in clinical medicine. Consistent with previously published data, we found that large changes in PVR (∼>50%) are necessary for RVFnRec ^10^. However, pulmonary compliance was significantly more predictive of RVFnRec than measures of steady load such as PVR and the composite measure of load, Ea. Studies have demonstrated that compliance has a more consistent relationship with outcome ^41-43^ and RV function ^44,45^ than does PVR. Also, experimental ^46^ and clinical ^47^ evidence indicates that compliance plays a significant role in maintaining optimal RV-PA coupling even at high PVR. Nevertheless, we found only a mild correlation between all standard load parameters and RV function. This finding may indicate that some afterload parameters not measured, like wave reflection ^48^, or load-independent factors, like possible direct prostacyclin-mediated effects on the RV such as reduced myocardial inflammation ^49^ or coronary microcirculatory effects ^50^, may play a role in RVFnRec. It is possible that RVFnRec may represent a more comprehensive therapeutic target, encompassing both load dependent and independent effects, than targeting load (i.e., PVR) alone.

We found that resting therapeutic improvements in afterload dissipate quickly during exercise as high PAP relative to cardiac output is seen even in subjects with RVFnRec. Likely, the drop in load seen at rest is negated at exercise as less affected pulmonary vessels meet maximal recruitment and distension as evidenced by very low α distensibility and high mPAP-C.O. slope in both groups. This observation suggests that pulmonary vascular remodeling is still likely advanced in RVFnRec despite such significant improvements in resting measurements of afterload. The demands on the RV by high afterload at exercise may explain why RVFnRec subjects maintain nearly twice the relative RV wall thickness (mass/volume) to that of controls despite near normalization of ventricular volume.

Our study has several limitations. It is a single center PAH cohort and therefore the results require validation amongst a larger population and preferably multiple centers. Validation is particularly important given that our ROC cutoff was assessed only at two time points (baseline and follow-up) and therefore does not account for intrinsic variability during the course of disease. For example, the RVEDV and Ca cutoffs may only be applicable to subjects with advanced PAH. Also, our study assessed VO2_peak_ only at follow-up and not the change in VO2 from diagnosis. However, the change in 6-minute walk distance from baseline to follow-up confirms a substantial difference in functional capacity between RVfnRec groups. Also, the feasibility of MRI is difficult at some centers where echocardiography is preferable. Future studies should validate our findings against echocardiographic metrics such as those recently proposed ^7^.

## Conclusions

Our data suggest that defining RVFnRec as a therapeutic normative -15 mL drop in RVEDV appears to be clinically useful. Among standard measures of afterload, an increase in Ca of 0.8 mL/mmHg by follow-up best predicts RVFnRec although factors outside afterload alone may be at play. We have shown that our definition of RVFnRec corresponds with near normalization of RV volume, but relative wall thickness remains high. The high relative wall thickness may be needed for output reserve in the face of high afterload at exercise.

## Data Availability

All data is available upon reasonable request from the contact author.

## Abbreviations

ANOVA: analysis of variance
BNP: brain natriuretic peptide
Ca: pulmonary vascular compliance
CI: confidence interval
CMR: cardiac MRI
CTEPH: chronic thromboembolic pulmonary hypertension
EDV: end-diastolic volume
EF: ejection fraction
ERS/ESC: European Respiratory Society/European Society of Cardiology
ESV: end-systolic volume
Ea: effective pulmonary arterial elastance
LV: Left ventricle
PA: pulmonary artery
PAH: pulmonary arterial hypertension
PAP: pulmonary artery pressure
PCWP: pulmonary capillary wedge pressure
PVR: pulmonary vascular resistance
RAP: right atrial pressure
REVEAL: United States Registry to Evaluate Early and Long-Term PAH Disease Management
RV: right ventricle
RVFnRec: RV functional recovery
sRVP: systolic RV pressure
SV: stroke volume
PTE: thromboendarterectomy
WSPH: World Symposium of Pulmonary Hypertension
VO2_peak_: peak oxygen consumption

## Acknowledgements

We would like to thank Andrew Swift (Department of Infection, Immunity and Cardiovascular Disease, University of Sheffield, Western Bank, Sheffield, UK) for sharing his CMR predicted reference equations with us. We would like to graciously acknowledge the assistance of the UAHD Biorepository at the University of Arizona. We would also like to acknowledge the many PH subjects of the UA registry/biorepository, the PVDOMICS study, and coordinators who have dedicated significant effort in the academic pursuit to further our field.

## Author Contributions

F.P.R., R.B., D.H.K., T.A., W.W.T.: study design; F.P.R., M.I., S.K.: patient recruitment, care and follow-up; MRI core interpretation; A.K., M.M.P., T.A., D.H.K.: rest and exercise hemodynamic core interpretation; F.P.R., M.I., S.K., J.D.W., W.W.T. .: data collection, maintenance, and analysis; R.J.B., F.P.R., R.P.F., P.M.H., A.R.H, E.M.H., J.A.L., E.B.: statistical analysis; F.P.R, A.B.L.:. drafted the original manuscript; F.P.R., R.N., R.R.V., R.B., R.J.B., A.B.L., D.H.K, T.A., critical revision of the manuscript for important intellectual content; F.P.R., M.I., S.K., G.J.B., N.S.H., R.P.F., P.M.H., A.R.H., E.M.H., J.G.G., J.A.L., E.B., S.C.E : principal investigator, had access to the study data and takes full responsibility for the integrity and accuracy of the data.

