## Supplemental methods, tables, and figures for "Classification and Predictors of Right Ventricular Functional Recovery in Pulmonary Arterial Hypertension"

**Online Data Supplement**

Franz P. Rischard*[1], Roberto J. Bernardo[2], Rebecca R. Vanderpool[3], Deborah H. Kwon[4], Tushar Acharya[5], Margaret M Park[4], Austin Katrynuik[6], Michael Insel[1], Saad Kubba[5], Roberto Badagliacca[7], A Brett Larive[8], Robert Naeije[9], Gerald J Beck[8], Serpil C Erzurum[10], Robert P Frantz[11], Paul M Hassoun[12], Anna R Hemnes[13], Nicholas S Hill[14], Evelyn M Horn[15], Joe G.N. Garcia[6], Jane A Leopold[16], Erika Berman Rosenzweig[17], W.H. Wilson Tang[4], Jennifer D. Wilcox[18]

*Dr. Rischard is both the first and lead author. Co-authors are then placed in order of contribution. If authors contributed equally, they are then listed alphabetically

[1]Division of Pulmonary, Allergy, Critical Care and Sleep Medicine, University of Arizona
[2]Division of Pulmonary, Critical Care and Sleep Medicine, University of Oklahoma Health Sciences Center, Oklahoma City, OK
[3]Division of Cardiovascular Medicine, The Ohio State University
[4]Department of Cardiovascular Medicine, Cleveland Clinic
[5]Divison of Cardiology, University of Arizona, Tucson, AZ
[6]Department of Medicine, University of Arizona
[7]Department of Cardiovascular and Respiratory Science, Sapienza University of Rome, Rome, Italy
[8]Department of Quantitative Health Sciences, Cleveland Clinic
[9]Department of Pathophysiology, Free University of Brussels, Brussels, Belgium
[10]Lerner Research Institute, Cleveland Clinic
[11]Department of Cardiovascular Medicine, Mayo Clinic
[12]Division of Pulmonary and Critical Care Medicine, Johns Hopkins University
[13]Division of Allergy, Pulmonary and Critical Care Medicine, Vanderbilt University Medical Center
[14]Division of Pulmonary, Critical Care, and Sleep Medicine, Tufts Medical Center
[15]Perkin Heart Failure Center, Division of Cardiology, Weill Cornell Medicine
[16]Division of Cardiovascular Medicine, Brigham and Women's Hospital, Harvard Medical School
[17]Department of Pediatrics and Medicine, Columbia University
[18]Department of Cardiovascular and Metabolic Sciences, Cleveland Clinic

**Supplemental Methods on Prospective Subject Follow-up Protocol**

All treatment-naïve subjects at the University of Arizona are encouraged to enroll in our protocolized PAH registry. Over 90% of incident patients participate at UA. This study began enrollment in 1/1/12. Enrolled subjects undergo right heart catheterization, 6 minute-walk test, echocardiography, cardiac MRI, BNP or NT pro-BNP at baseline (diagnosis for most). Invasive cardiopulmonary exercise (iCPET) testing is done for patients' functional class (FC) I-IIIB. This testing is repeated on therapy at 3-6 months and 6-12 months depending on baseline presentation and achievement of therapeutic goals (see therapeutic strategy). All patients not FC IV undergo iCPET at follow-up.

**Supplemental Methods on Therapeutic Strategy**

Subjects were categorized by therapeutic strategy based on the European Respiratory Society/European Society of Cardiology (ERS/ESC) guidelines of 2015 at which goal-directed mono or sequential therapy was replaced by up-front combination therapy [1]. Prior to 2015, a goal-directed sequential combination therapy was used based on achievement of functional class (FC) I-II. After diagnosis, if FC I-II was not achieved by 3-6 months re-assessment, then combination therapy was added. Therefore, if a patient remained on treprostinil monotherapy, the therapeutic goal was achieved. In this context, combination therapy before 2015 represents treatment failure of treprostinil monotherapy. After 2015, all patients were placed on up-front combination therapy in accordance with updated guidelines.

**Supplemental MRI Methods**

The imaging protocol included short axis (SAX) stack analysis of volumes for both the left ventricle (LV) and right ventricle (RV). Q flow analysis included phase imaging of the main pulmonary artery (PA). Additionally, PA Q flow analysis and SAX data were used to calculate TV regurgitant volume (TVRV) as (RV stroke volume (RVSV) – PA Q flow forward volume), and regurgitant fraction as (TVRV / RVSV).

RV and left ventricular (LV) ejection fraction (EF), RV and LV end-diastolic/systolic volumes (EDV/ESV), RV stroke volume (SV), RV and LV mass were derived by contouring the endocardial border on the end-diastolic phase and the endocardial border on the end-systolic phases with papillary muscles and trabeculations included as part of the blood pool. RV and LV end systolic phases and end diastolic phases were identified on separate phases when significant RV/LV uncoupling was present and was defined as the phase where the RV or LV was the smallest to define end systole and largest to define end diastole. The long axis 4 chamber view was used as a reference image allowing for determination of basal slice inclusion while outlining the LV and RV endocardial border. RV and LV stroke volumes were compared and used to additionally guide inclusion of a basal slice (only when there was no suspicion of shunt). Careful assessment to delineate the RVOT from the pulmonic valve and right atrium was performed by stepping through each cardiac phase multiple times to identify the appropriate borders. The basal-most left ventricular slice was included if the myocardium extended to ≥ 50% of the circumference of the short axis slice.

RV mass was determined on the end-diastolic phase by contouring the epicardial border in addition to the endocardial border. Contour smoothing was performed on the endocardium and trabeculations included in the blood pool (excluded from mass calculations). The interventricular septum was included with the left ventricular mass. The myocardial volume for each slice was calculated by multiplying the area of the RV wall by the slice thickness. RV and LV mass was calculated as the product of the sum total of the myocardial slice volumes for each ventricle multiplied by 1.05 g/cm$^3$ [2]. Ventricular volumes and ventricular mass were indexed by body surface area.

An intra-class coefficient was calculated using a two-way random effects model detecting an absolute difference in ventricular volumes to assess inter-reader variability. CMR inter-reader variability was examined by intra-class correlation performed on 50 subjects (22 controls and 28 PAH) demonstrating high agreement of 0.98 (95% CI 0.98-0.99, $P<0.001$), 0.99 (95% CI 0.98-0.99, $P<0.001$), and 0.96 (95% CI 0.92-0.98, $P<0.001$) for RVEDV, RVESV, RVEF, respectively.

### Supplemental Tables

| MRI Variables | $VO2_{peak}$ >15 mL/kg/min | | |
|---|---|---|---|
| | Youden Index (Cutoff) | AOC (95%CI) | P-value |
| **RVEF** | | | |
| Change in RVEF (%) | 0.51 (4.1) | 0.74 (0.66-0.89) | 0.0001 |
| Relative change in RVEF (%) | 0.61 (15) | 0.74 (0.62-0.86) | 0.002 |
| RVEF at FU (%) | 0.55 (37) | 0.73 (0.62-0.88) | 0.001 |
| **RVEDV** | | | |
| Change in RVEDV (mL) | 0.74 (-15) | 0.87 (0.77-0.96) | 0.0001 |
| Relative change in RVEDV (%) | 0.72 (-6.5) | 0.87 (0.77-0.96) | 0.0001 |
| RVEDVI at FU (mL/m2) | 0.40 (112) | 0.67 (0.53-0.80) | 0.03 |
| **RVESV** | | | |
| Change in RVESV (mL) | 0.63 (-47) | 0.86 (0.77-0.95) | 0.0001 |
| Relative change in RVESV (%) | 0.72 (-30) | 0.88 (0.80-0.98) | 0.0001 |
| RVESVI at FU (mL/m2) | 0.42 (66) | 0.72 (0.59-0.86) | 0.004 |

**Table E1. Potential right ventricular defining MRI parameters of functional recovery predicting exercise capacity at follow-up.** Exercise capacity is defined by peak oxygen consumption >15 mL/kg/min by guidelines [3] and evidence [4]. RVEF, right ventricular ejection fraction; RVEDV, RV end-diastolic volume; RVESV, RV end-systolic volume; FU, follow-up.

| | PAH | | | Healthy Control | | |
|---|---|---|---|---|---|---|
| | Mean±SD | 95% CI | 5/95 Percentiles | Mean±SD | 95% CI | 5/95 Percentiles |
| **RVEDV (%)** | **116.4±41.0*** | 105.8-126.9 | 73.4/224.1 | 71.1±13.0 | 67.8-74.4 | 51.0/93.4 |
| **RVEDVI (%)** | **151.9±60.2*** | 136.8-168.3 | 91.2/268.6 | 90.8±16.8 | 86.4-95.0 | 65.8/120.6 |
| **RVESVI (%)** | **298.9±155.5*** | 187.1-980.0 | 188.4/768.7 | 114.0±29. | 106.4- | 67.9/171.5 |
| **RVEF (%)** | **52.72±14.8*** | 23.8-84.9 | 29.9/71.6 | 86.99±9.4 | 84.6-89.4 | 71.1/101.4 |
| **RVEF MESA (%)** | **51.1±14.6*** | 47.5-55.2 | 22.2/78.0 | 84.2±8.7 | 82.0-86.4 | 70.2/98.7 |
| **RVMI (%)** | **154.3±67.6*** | 59.0-339.4 | 98.0/279.8 | 43.55±13. | 40.1-46.8 | 23.4/71.6 |
| **LVESVI (%)** | 114.5±41.0 | 15.4-246.8 | 71.7/205.1 | 117.0±25. | 110.6- | 79.0/173.1 |
| **LVEDVI (%)** | 86.7±21.0 | 60.4-123.9 | 66.5/110.6 | 92.6±16.1 | 88.5-96.7 | 67.1/117.1 |
| **LVEF (%)** | 84.9±12.1 | 35.3-124.6 | 45.7/106.2 | 86.9±6.1 | 85.4-88.5 | 76.1/98.0 |
| **LVMI (%)** | 67.6±30.2 | 81.0-105.4 | 85.0/100.9 | 66.0±15.3 | 62.0-69.9 | 42.9/98.0 |

**Table E2. Percent predicted cardiac MRI ventricular volumes, function, and mass by PAH and healthy control cohorts.** Predicted values are obtained from the MESA cohort equations [5] and ref.[6]. RVEDVI, right ventricular end-diastolic volume index; RVESVI, right ventricular end-systolic volume index; RVEF, right ventricular ejection; RVMI, right ventricular mass index; LVEDVI, left ventricular end-diastolic volume index; LVESVI, left ventricular end-systolic volume index; LVEF, left ventricular ejection-fraction; LVMI, left ventricular mass index. *Indicates P<0.05 versus Healthy Control subjects. Percentiles are based on the weighted average.

**Supplemental Figures**

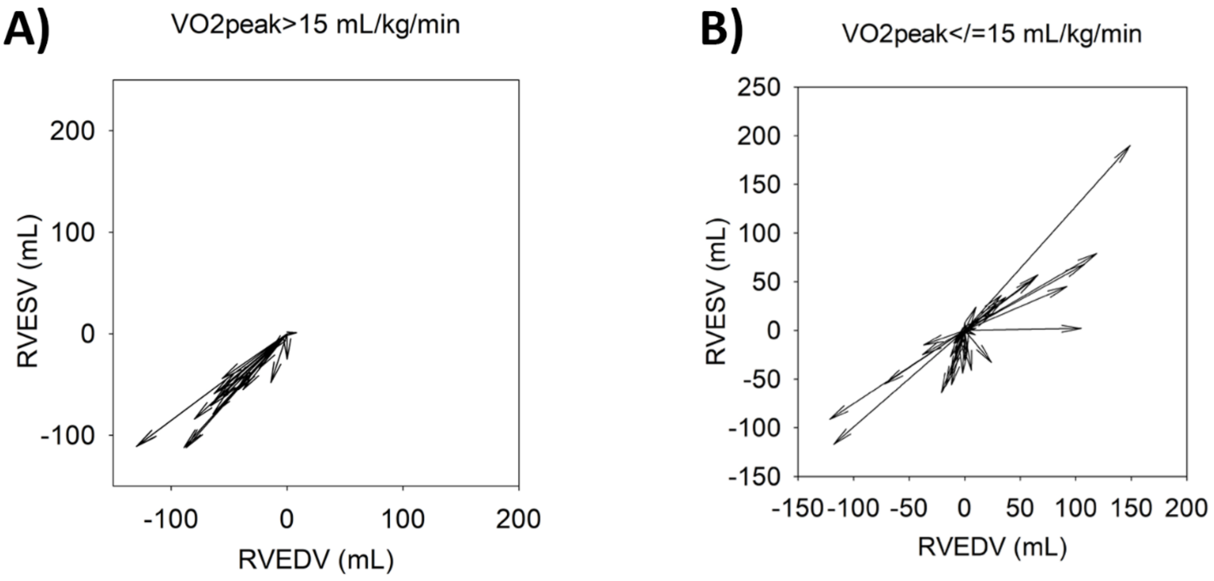


**Figure E1. Vector Plots demonstrating changes in RV end-systolic (RVESV) and diastolic (RVEDV) volumes from baseline to follow-up by exercise capacity peak oxygen consumption ($VO2_{peak}$) > or ≤ 15 mL/kg/min.** The high exercise capacity group (A) have a reduction in both ESV and EDV whereas some of the low exercise cohort have a reduction in ESV without a change in EDV (B). Therefore, a drop in RVEDV is necessary for high exercise capacity.

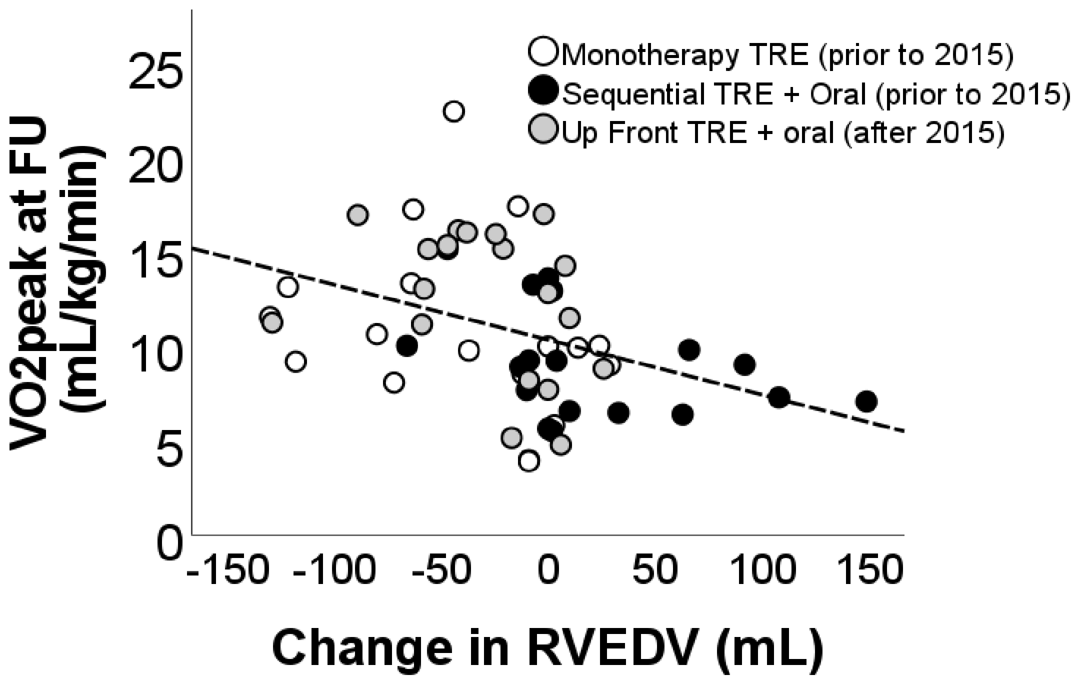


**Figure E2. The effect of therapeutic approach on change in RVEDV and VO2peak at follow-up.** Up-front combination therapy was associated with odds-ratio of 10.4 (CI 1.9-56.6, P=0.007) of RVFnRec relative to goal-directed sequential combination therapy. See supplemental methods for therapeutic strategy.

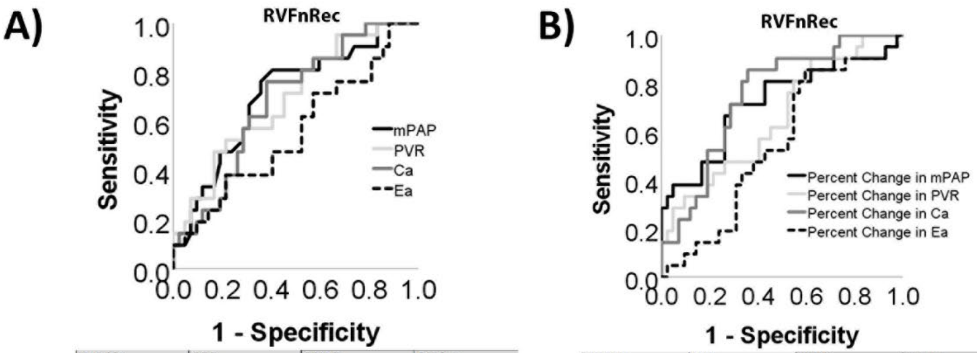


| Variable | AOC | 95% CI | P Value |
|---|---|---|---|
| mPAP at FU | .705 | .56-.84 | .008 |
| PVR at FU | .693 | .56-.82 | .013 |
| Ca at FU | .683 | .55-.82 | .019 |
| Ea at FU | .556 | .40-.71 | .475 |

| Variable | AOC | 95% CI | P Value |
|---|---|---|---|
| Change in mPAP | .72 | .58±.87 | .004 |
| Change in PVR | .66 | .52±.80 | .038 |
| Change in Ca | .75 | .63±.87 | .001 |
| Change in Ea | .55 | .41±.70 | .08 |

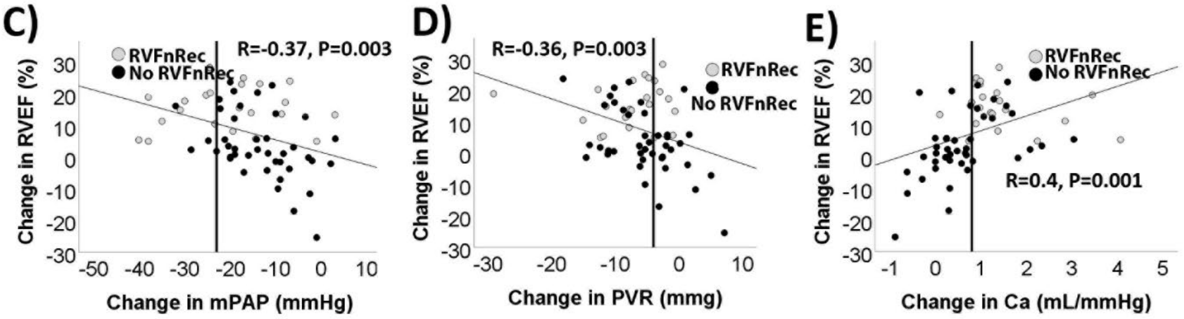


<u>**Figure E3.**</u> **Afterload changes predicting right ventricular functional recovery**. Receiver operating curve analysis predicting RVFnRec of absolute values at follow-up (FU) (**A**) and relative longitudinal changes from baseline to follow-up (**B**) in mean pulmonary artery pressure (mPAP), PVR, Ca, and effective pulmonary elastance (Ea). Changes in all afterload parameters were mildly correlated changes in RV ejection fraction (RVEF) (**C-E**).

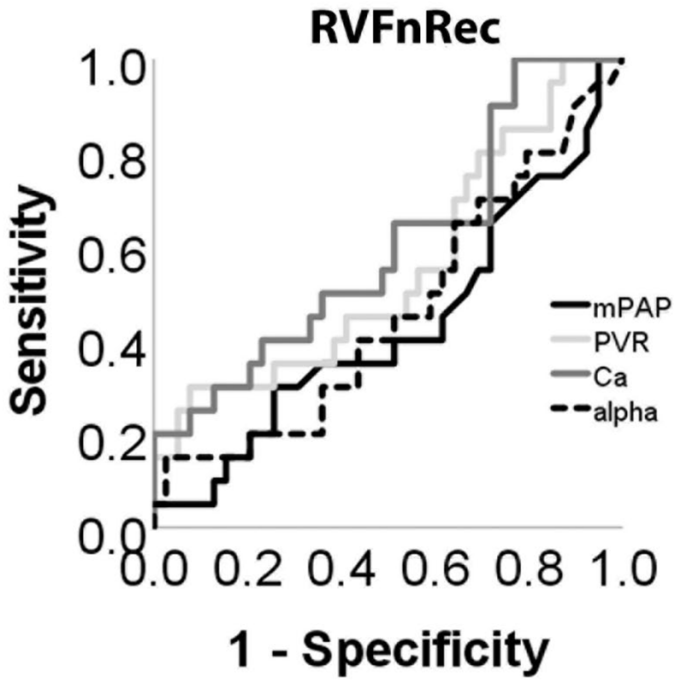


| Variable | AOC | 95% CI | P Value |
| --- | --- | --- | --- |
| Change in mPAP | .43 | .27±.59 | .38 |
| Change in PVR | .55 | .39±.72 | .50 |
| Change in Ca | .60 | .45±.77 | .21 |
| Change in Ea | .46 | .30±.62 | .63 |

**Figure E4. Receiver operating curve analysis of exercise afterload parameters predicted right ventricular functional recovery at follow-up**. At exercise, afterload parameters loose accuracy in predicting RVFnRec.
